# Community Detection and Patient Experience Analysis in Reddit Conversations on Janus Kinase Inhibitors using Large Language Models

**DOI:** 10.64898/2026.02.02.26345429

**Authors:** Tawakalit Omolabake Agboola, Shumyl Akbar, Udoka Duruaku, Ali Al-Janabi, Adriana Ioannou, Jia Hui Megan Loh, Charlene Murphy, Zenas Z. N. Yiu, Oluwaseun Ajao

**Affiliations:** School of Computing and Mathematics, Manchester Metropolitan University, Manchester, UK; Division of Musculoskeletal and Dermatological Sciences, Manchester NIHR Biomedical Research Centre, Manchester Academic Health Science Centre, University of Manchester, UK; National Eczema Society (NES), 82 Tanner Street, London, UK; Dermatology Centre, Northern Care Alliance NHS Foundation Trust UK

## Abstract

The emergence of Janus kinase (JAK) inhibitors, a relatively new class of medications for autoimmune and inflammatory conditions, has been accompanied by reports of adverse effects observed during clinical trials. However, uncertainty over their safety and efficacy in wider, unselected populations has led to discussion and speculation on social media such as Reddit. Social networks represent a novel, rich source of real-world pharmacovigilance data. They are also an environment where unverified information about these medications may circulate. This paper analyzes Reddit conversations related to JAK inhibitors, applying graph modeling and community detection techniques using Neo4j and the Louvain algorithm. Data from 2011 to 2024 were collected, cleaned, and used to construct a directed graph, incorporating posts, comments, users, and drug mentions as nodes and their interactions as edges. Advanced computational methods, including large language models, were utilized to analyze textual data and identify patient-reported experiences that diverge from current medical consensus. This study systematically maps online discourse and identifies key participants to understand how patient experiences and concerns about JAK inhibitors are shared within communities.

The findings show that various subreddits serve as hubs of information in which key influencers are spreading both positive and negative information within the Reddit ecosystem. Highlighting the potential to integrate graph-based approaches, Neo4j, and advanced LLMs in real-time pharmacovigilance, this study presents compelling evidence of the emerging conversations surrounding JAK inhibitors and how they affect public health.

**Author Summary:** People often turn to Reddit to share their experiences with medications, including Janus kinase (JAK) inhibitors, which are used to treat autoimmune conditions such as arthritis, eczema, and alopecia areata. These drugs are fairly recent, and some safety concerns have been identified, making discussions about them on the internet a mixture of personal stories, questions, and statements which may not correspond to serious or established medical literature. [60]

In this research we analyzed over ten years of Reddit discussions to explore how individuals discuss JAK inhibitors and how both correct and possibly misleading information circulates within groups. We integrated graph-based techniques, which illustrate the connections among users and conversations with AI tools that identify claims at odds, with clinical guidelines.

We applied the term ”divergent patient experiences” exclusively to comments that contradict regulation or evidence-based sources, while personal accounts and feelings of individuals are not classified as divergent patient experiences.

Our findings demonstrate that a very small number of users initiate a large proportion of conversations and discussions tend to revolve around the main health topics. This approach of using social media to monitor public health opinions shows the manner in which it avails information regarding real patient concerns, but it also shows the requirement of expert supervision when using AI to appraise health information being shared online.

## 1 Introduction

Janus kinase (JAK) inhibitors are an important class of medications for the treatment of autoimmune and inflammatory diseases, including rheumatoid arthritis, inflammatory bowel disease, atopic dermatitis, and alopecia areata [11] [54]. These work by inhibiting the JAK-STAT signaling pathway which helps reduce inflammation [29]. However, patient concerns have been raised about their long-term safety profile [54]. There is evidence of an association between JAK inhibitors and serious adverse events (AEs), especially cardiovascular AEs and malignancy as demonstrated by the ORAL Surveillance trial, and regulatory black box warnings for these drugs have been filed [12, 54]. These warnings have been based on data from trials in people with rheumatoid arthritis, and it is unclear whether the same risk is present when people with other conditions are prescribed JAK inhibitors [35, 56]. This underscores the need for information on the risk of these medicines from large real-world populations, including patient perceptions and experiences with these medications [6, 51].

Social media platforms, such as Reddit, have become a repository of patient reports of outcomes, experiences on various treatments, efficacy, and safety [6, 51]. These platforms have offered unprecedented opportunities for real-time pharmacovigilance [6, 58]. There are challenges with using these data for analysis, particularly around identifying novel safety signals amid varied patient experiences, which can cloud public understanding and influence health decisions [10, 13]. Vosoughi [59] showed that false information flows significantly farther, faster, deeper, and more broadly than true facts on social networks emphasizing the importance of developing ways to identify and analyze patterns in patient-reported experiences in such settings. These conversations can be studied using community detection to show the organic clusters of users centered around common interests [17, 21, 47].

Within Reddit, discussions about JAK inhibitors do not occur in isolation but take place in shared communities that focus on different aspects of the drugs [20, 39]. These communities (known as subreddit in the Reddit platform) serve as echo chambers where relevant narratives are magnified and can provide a more refined view of public sentiment and emerging safety concerns about these drugs. Community detection and key participant analysis propagating this information provide structural insight into the mechanics of how discussions unfold, but understanding the content of the conversation is also important [10, 18].

As patient-reported health experiences are complex in nature, tools that can analyze large volumes of text (such as thousands of Reddit posts and comments) and identify inconsistencies between user-generated content and established medical knowledge are needed [10, 49]. In this study, Large Language Models (LLMs) are integrated to evaluate the credibility and reliability of the information shared within these Reddit communities.

The main objectives of this study were to:

- Identify and analyze unique communities: participating in discussions regarding JAK inhibitors on Reddit.
- Identify and analyze patient-reported experiences that diverge from current medical consensus, recognizing these may represent important pharmacovigilance signals, emerging adverse effects, or areas requiring enhanced patient communication.
- Identify key influencers: analyze the significance of influencers in these communities by using measures of centrality to gauge their influence on spreading information within the reddit platform.

## 2 Methodology

### 2.1 Data Collection

Reddit data were collected using the asyncpraw library to scrape posts and comments discussing JAK inhibitors from 2011-01-13 to 2024-09-22 [6] as shown in fig1. Data extraction focused on key medications and conditions associated with JAK inhibitors, using targeted keywords such as the combination of Janus Kinase Inhibitor (JAKi) drugs and the medical conditions they treated. All data collection was carried out according to the terms of service for Reddit’s API and user privacy was maintained [51]. No personally identifiable information was collected and data have been in compliance with General Data Protection Regulation [6]. Data was cleaned after collection to ensure consistent formatting. Text fields such as Comment Body and Title were standardized, transformed to lowercase, and whitespace and special characters were removed.

**Fig 1.**
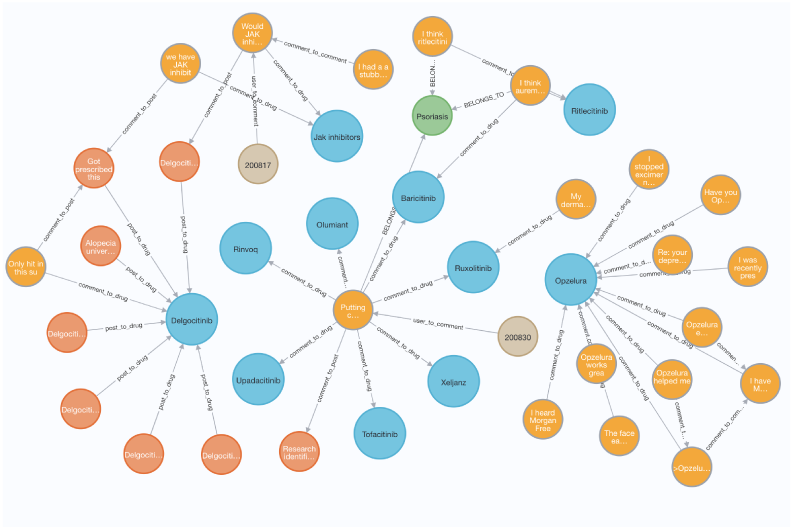
Sample Structure of the Community Graph

In addition, to preserve privacy and anonymity, all user identifiers, such as Reddit usernames, were pseudo-anonymised through a hashing process (SHA-256) [6]. The hashed values cannot be reversed, and thus users cannot be identified directly. The user identifiers have been changed to their hashed equivalent for the analysis and the original data were stored securely in a different database.

### 2.2 Graph Modelling

To analyze the structure and interactions of Reddit discussions related to JAK inhibitors, a directed graph was built using NetworkX, which is a Python-based library that provides network analysis components [19, 24]. In the graph, users, posts, comments, and drug mentions serve as nodes, whereas the interactions used to connect them serve as edges. This approach allows for a structured examination of how discussions evolve; sharing of patient experiences; and which users have a large influence in shaping discourse [20, 39]. GraphRAG (Retrieval-Augmented Generation on Graphs) complements traditional text processing techniques with the topological information provided by graph-based data, including user behaviors and comment threads, with the contextual understanding of large language models [16, 30, 46].

To build the graph, posts, comments, users, and drug mentions were mapped based on conversations. Each post was a node with links to the author who created it and other users’ comments about it. Comments were nested by being directly linked to the original post or to other comments, which create a hierarchical discussion thread. Each user was associated with the posts and comments they made, allowing us to analyze engagement trends. The drug mentions within posts and comments were detected and coupled to the relevant nodes in the discussion, allowing exploration of the frequency and context in which JAK inhibitors were mentioned.

During construction, the relationships between different entities were preserved. Popularity scores for posts and comments were included to check whether increased engagement resulted in propagation of divergent patient experiences. This dataset was cleaned to improve reliability by removing duplicate entries and adding only structured data fields such as post and comment IDs, timestamp, user interactions, and content.

### 2.3 Neo4j Integration

The Neo4j integration was a key step in structuring and analyzing the dataset to ensure that complex relationships between users, posts, comments, and mentions of drugs were preserved and efficient queries were possible. A graph-based model like Neo4j’s offered a scalable and natural way to represent social media data and a greater chance of gaining insights about the interactions that are happening in Reddit conversations [24, 46]. Using Neo4j querying functions, it was possible to explore relationships, fetch patterns of discourse, and visualize the spread of information across different communities [20].

To perform this operation, the Networkx structure was first exported into a suitable format for Neo4j. The dataset was organized into separate files such as nodes and edges, with all properties by entity type preserved. Each of the posts, comments, users, and mentions of drugs were kept as separate node types, to retain important metadata attributes, such as text, timestamp, authorship, and subreddit. In most social media platforms, an entity can be represented as nodes and relationships between these nodes, for example, user-post interactions, comment threads, drug mentions, and the relationships between these nodes were mapped, ensuring that the discussion hierarchy remained intact. Due to the size of the dataset it was also important to execute the import process in an efficient way to prevent computational bottlenecks, as there were over 248,450 nodes and 324,483 relationships.

Batch processing techniques were implemented, enabling incremental data insertion instead of processing the entire dataset at once to mitigate this issue. In addition to that, indexing strategies were employed for fundamental identifiers such as post and comment IDs, thereby allowing queries to be executed quickly without excessive computational overhead. With the complete dataset uploaded, Neo4j allowed an interactive environment, exploring the relationships within the discussion network. The graph structure mirrored how people interact on Reddit, a post is frequently at the center of a discussion, with comments branching off as response threads out of the main body. The users were also connected to the content they produced, enabling analysis of engagement behaviors and detection of opinion leaders. References to JAK inhibitors were explicitly linked back into the discussion of each drug, examining how the frequency of reference across different drugs related to the discussion context. The visualization in Fig(1) is a sample structure of neo4j where orange:comment, red:post, blue:drugs, brown:users.

### 2.4 Community Detection and Validation in Reddit Conversations

Understanding the structure of discussions surrounding JAK inhibitors on Reddit requires identifying natural groupings of users, posts, and comments that emerge within the social media discourse [17, 21]. These communities often govern the way experience sharings, as some clusters amplify misleading content, while others foster evidence-based discourse [13, 59]. In this study, community detection is applied to uncover these hidden structures using the Louvain algorithm, the most widely used method for optimizing modularity in large-scale networks [5]. The community is validated using statistical measures including modularity, conductance, and degree distribution analysis, to ensure robustness and interpretability of results [18].

The Louvain algorithm is a greedy optimization method designed to maximize modularity in large-scale networks. Detects densely connected subgroups (communities) by repeatedly reassigning nodes to communities in a way that maximizes the gain in modularity. It effectively solves large-scale graphs with complex community structure [5, 37]. Mathematically, for a graph G = (V, E), with V as the nodes (users, posts, comments, drugs) and E as the edges (interactions between these nodes), we define the modularity function Q such that:

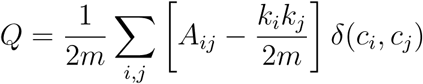

where;

*Aij*: represents the adjacency matrix of the graph,
*k_i_k_j_*: are the degrees of the nodes,
m is the total number of edges in the network, *δ*(*c_i_, c_j_*): is an indicator function, which is 1 if nodes i and j belong to the same community and 0 otherwise.

#### Modularity Score

As defined previously, modularity (Q) is the primary metric used to evaluate the quality of the community structure [17, 47]. Higher modularity indicates that the detected communities are densely connected internally, while weakly connected between the communities [47].

Empirical studies suggest that: *Q >* 0.3 suggests a significant community structure.

*Q >* 0.6 indicates a highly modular network, where communities are strongly self-contained. In simpler terms, modularity (Q) is a metric that describes how well the network separates into “communities” where each user has a higher degree of interaction with other members of the community than outsiders, with higher value indicating more distinct community boundaries [47].

#### Conductance

Conductance is a measure of how well-separated a community is from the rest of the network. It is defined as;

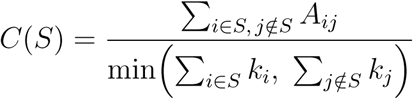

where:

*S*: represents a detected community, is the number of edges connecting nodes inside and outside the community,
*k_i_k_j_*: are the degrees of the nodes.

### Degree Distribution and Power-Law Analysis

The degree distribution of a network refers to how connections are distributed among nodes. Real-world social networks often have a power-law degree distribution, where a small number of nodes (influencers) have many connections and most nodes have only a few [3, 55]. The probability P (k) that a randomly selected node has degree k follows a power law:

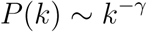

where *γ* is the power-law exponent.

In order to statistically test if the degree distribution follows a power-law, we apply a Kolmogorov-Smirnov (KS) test [36]. The statistic of the KS test D is given by: where:

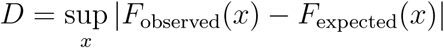

*F_observed_*(*x*) is the empirical cumulative distribution function (CDF). *F_expected_*(*x*) is the theoretical CDF under the power-law model. In this study, the KS test produced *D* = 0.47*, p <* 0.0001, further confirming that the degree distribution is significantly different from a random network. The estimated power-law exponent *γ* = 2.11, suggesting a scale-free structure characterized by a small number of influential users dominates the discussion.

To further compare different statistical models (power-law, log-normal, and exponential) [8], the Akaike Information Criterion (AIC) is used:

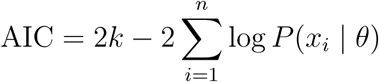

where: *k* is the number of model parameters, is the likelihood of the observed data under the model.

### 2.5 GraphRAG Integration for Patient Experience and Signal Analysis

Patient-reported experiences on social media offer valuable pharmacovigilance data but requires sophisticated methodologies that capture content at a deeper level than simple keyword-based detection [6, 51]. GraphRAG (Retrieval-Augmented Generation on Graphs) combined with Neo4j graph database and pre-trained LLMs enables contextualizing, labeling, and analyzing Reddit conversations about JAK inhibitors in a novel approach defined in this study [16, 30, 46]. As discussed in previous sections (2.2, 2.3 and 2.4), the Neo4j database was organized as a directed graph, aimed at maintaining the complicated interconnections between users, posts, comments, and mentions of drugs. To leverage this graph for advanced patient experience analysis and community labeling, a retrieval-augmented generation framework, GraphRAG was employed. This approach incorporates structural advantages from graph-based methods and contextual nuances from LLMs in order to provide a context-based exploration of the detected communities of textual content. The first part of this process involved exporting the Neo4j graph into a format that is compatible with GraphRAG, all nodes (such as, posts, comments) and their properties (such as, text, timestamps, authorship) were preserved. Using the create vector index function, vector indexes for both posts and comments were created, with embeddings produced in 1536 dimensions and using cosine similarity as the distance metric. Such indexes allowed fast retrieval of relevant content using semantic similarity, an important aspect for the following analysis of patient experiences. In this study, the OpenAI pretrained model GPT-3.5-turbo was chosen particularly for its significant natural language understanding competence and contextual nuance, especially across a wide range of textual forms, and successful performance in related tasks such as detection of divergent patient experiences and semantic analysis as illustrated in previous works [10, 49]. The large-scale unstructured social media content with context-specific nuance made it a fit to evaluate the Reddit-based discussions. To guide the identification of divergent patient experiences, we developed structured prompts based on medical standards and pharmacovigilance guidelines. These prompts instructed the model to flag content as potentially divergent from current evidence only if it exhibited characteristics such as:

i. exaggerated or unverified medical claims,
ii. statements that contradicted regulatory-approved guidance (FDA),
iii. promotion of off-label use without evidence,
iv. commercial endorsements lacking disclosure

Moreover, its accessibility through API, and fine-tuning suitability, was compatible with the study’s real-time pharmacovigilance aims, addressing scalability at no cost to accuracy.

Embedding the graph data involved transforming textual content into dense vector representations using an OpenAI embedding model (text-embedding-ada-002). This approach is also based on the EmbeddingGenerator class, which takes batches of nodes and gets a node embedding, which allows it to scale to the size of the dataset and not add additional computational overhead. Traditionally, each post and comment were embedded separately and an embedding property was assigned to each node in Neo4j. These generated embeddings were able to capture such semantic relationships within the entire discourse, allowing GraphRAG to retrieve contextual nodes pertaining to each respective query. A MultiIndexRetriever was implemented to name the identified communities (identified by Louvain algorithm, Q=0.9156) of detected communities. This retriever aggregated results from both post and comment indexes, ranking them by similarity scores and selecting the top k=3 results. The LLM then examined the retrieved discussions to assign short keyword-based labels (such as, “Alopecia treatments,” “Eczema”) and summaries.

Utilizing the GraphRAG module, the neo4j was connected to the LLM by executing the GraphRAG pipeline using a customized class called GraphRAGAnalyzer. The retriever was initialized, the nodes were embedded, and LLM was queried to generate labels that were written back to the neo4j database as spring nodes with a communityLabel property. This integration not only structured the unstructured social media data, but also computed a context for the topics of interest by putting the discussions in the context of labeled communities, thus setting the stage for detection of divergent patient experiences. The approach aligns with the aim of the study to combine graph-based methods with LLMs, improving pharmacovigilance in real time as much as possible by identifying discourse patterns and influential narratives.

### 2.6 Analysis of Patient-Reported Experiences

Following the integration of GraphRAG with the Neo4j graph database and the labeling of communities, the framework was extended to identify content that diverges from established medical consensus on the Reddit discussions around Janus kinase (JAK) inhibitors [10, 49]. Posts were flagged as potentially divergent from current evidence when the model estimated that their content may diverge from established regulatory guidance or clinical evidence for instance, exaggerating efficacy (”miracle cure”), downplaying known or potential risks (such as malignancies), or promoting unverified off-label use. Using a customized retrieval and evaluation method, the analysis was conducted on the top five communities based on posts and comments volume. The GraphRAG pipeline was implemented through the GraphRAGAnalyzer class, which retrieved pertinent content from vector indexes consisting of posts and comments populated with 1536-dimensional embeddings and guided by cosine similarity. A Multi-IndexRetriever was used to fetch posts and their respective comments based on the relationship between comments and posts, allowing for an in-depth analysis of discussion threads. This approach builds on the community detection framework, shifting the focus from structural analysis to content credibility within these social hubs.

To compute divergence likelihood scores, GPT-3.5-turbo was prompted to assign a likelihood score (0-1) per sentence. The overall divergent patient experiences confidence score is determined through combining two important factors against each post, how misleading each post is (divergence score) and the degree of relevance of the post concerning the search query (retrieval relevance score). GPT-3.5-turbo generated estimated likelihood scores, where higher values indicate a greater model-estimated probability that content may be misleading. The most misleading sentence and the average of all sentence scores is used as the post’s overall divergence score. Since not all retrieved posts were equally relevant to the query, a relevance score (calculated using cosine similarity) determined how strongly each post influenced the final divergence score. These scores were aggregated using:

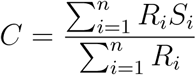

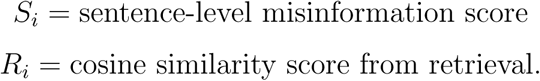

A cosine similarity threshold (≥ 0.5) was used to maintain the consistency of the responses. The LLM scores do not represent explicit probabilistic truth values, but do provide a structured proxy for the likelihood of divergence from current evidence in the dataset.

The PageRank metric, used to assess user influence [47], is defined mathematically as:

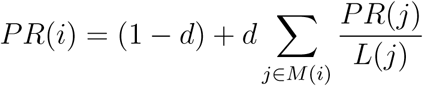

where

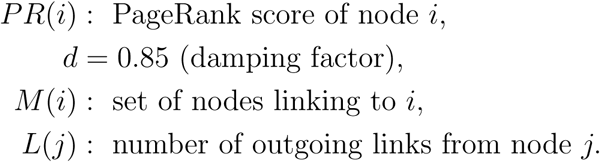

## 3 Result

### 3.1 Reddit Data Overview

Analyzing Reddit conversations about Janus Kinase (JAK) inhibitors from 2011 to 2024 provides valuable insights into the evolving landscape of online health discussions [6]. Over this 13-year period, the dataset reflects evolving community structures, the proliferation of divergent patient experiences [10, 13], and the influential role of key users in shaping public perception [20, 39]. By leveraging advanced graph-based methodologies and large language models (LLMs) [7, 24], we identified critical patterns in how discussions on JAK inhibitors unfold across Reddit communities.

A clear trend of increasingly active engagement is visible in Fig(2, where the number of comments surpass the number of original posts. This divergence became particularly evident, indicating increased public interest in JAK inhibitors due to regulatory events and rising awareness of autoimmune conditions [2].

**Fig 2.**
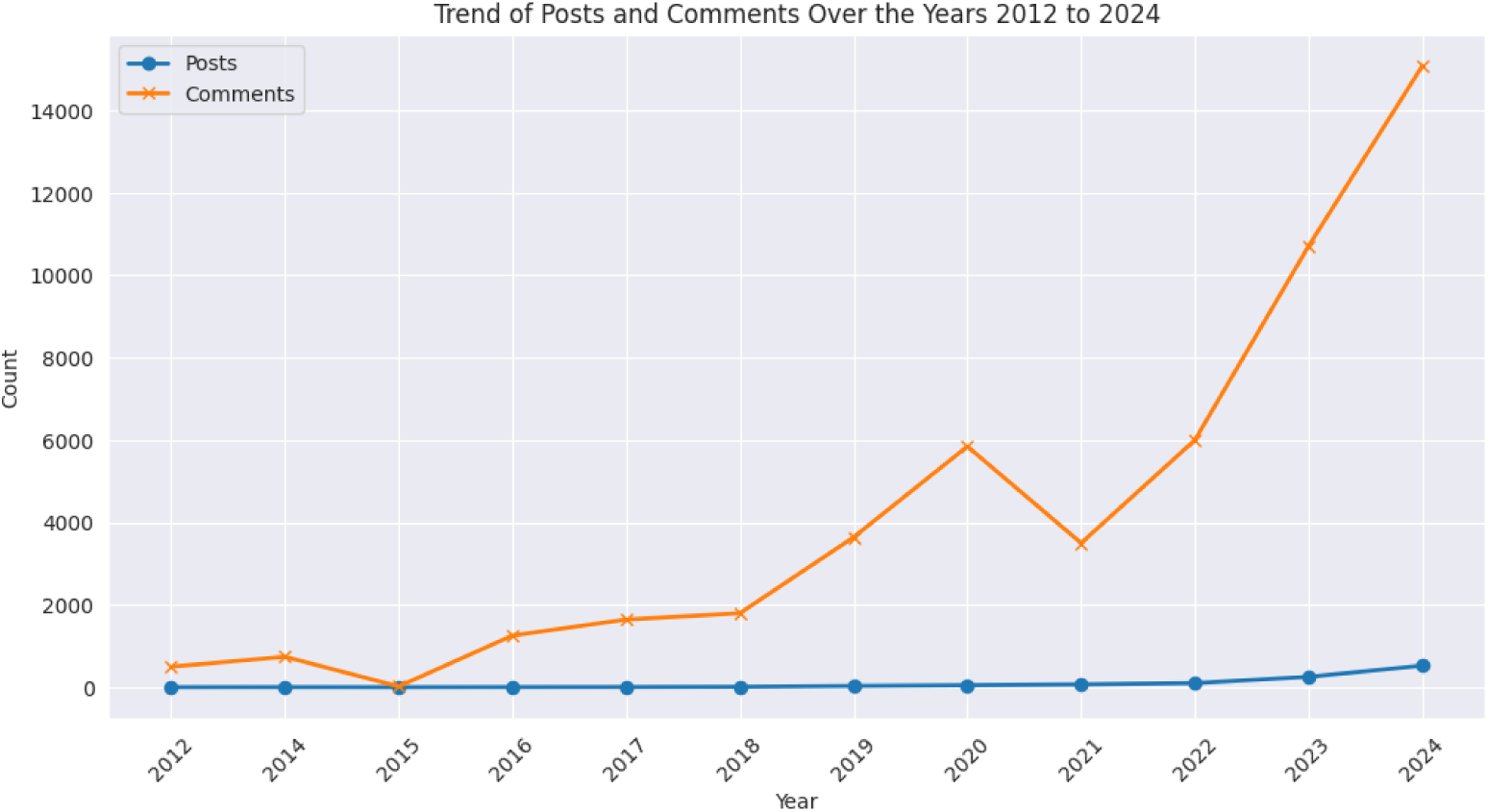
Trend of Post and Comment over the years 2011 to 2024

### 3.2 Graph Modelling and Community Detection

The examination of Reddit conversations about Janus Kinase (JAK) inhibitors from 2011 to 2024 revealed intricate community structures, the prevalence of divergent patient experiences, and the significant influence of key users within these online discussions. A directed network was constructed with 248,450 nodes (users, posts, comments, and drug mentions) and 324,483 edges, where edges represented the interactions between these entities. Using the Louvain algorithm on this graph resulted in 337 communities with a modularity score of 0.9156 [5, 47], which indicates that the communities are tightly knit and focused on specific themes. Conductance (0.0396) suggested good isolation [18], and the degree distribution followed a power-law (exponent *γ* = −2.11), confirmed by a Kolmogorov-Smirnov test (*D* = 0.47*, p <* 0.0001) [3, 36, 55].

**Table 1.** Community Detection Metrics Results from the Reddit Conversations on JAK Inhibitors.

| Metric | Result | Interpretation |
| --- | --- | --- |
| Modularity ($Q$) | 0.9156 | Indicates a highly modular network with strongly self-contained communities ($Q > 0.6$). |
| Conductance ($C(S)$) | 0.0396 | Measures inter-community connectivity; lower values suggest better isolation. |
| Power-law exponent ($\gamma$) | -2.11 | Suggests a scale-free network where a few nodes (influencers) dominate connections. |
| Kolmogorov-Smirnov ($D$) | 0.47, $p < 0.0001$ | Confirms the degree distribution significantly follows a power-law, rather than a random network. |

Among the models tested, the power-law model had the lowest AIC score (-2985961.23), meaning that it is the best-fitting model to describe how influence is distributed among users in Reddit discussions indicating a scale-free network where a few influential users dominated discussions. The top five communities represented by ID numbers 46573 (16,380 nodes), 130550 (13,301 nodes), 92085 (11,055 nodes), 92479 (11,017 nodes), and 40201 (5,717 nodes) accounted for approximately 31.5% of the overall nodes, indicating significance as hubs of activity in the network.

**Table 3.**
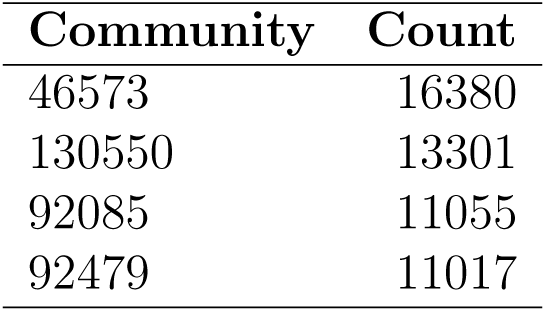
Top 5 Communities and Their Counts.

GraphRAG was applied to assign thematic labels to these communities, such as ”Alopecia treatments”, ”Psoriasis”, and ”Eczema”, which correspond to the primary indications of JAK inhibitors [16, 30, 46].

Analysis of Patient-Reported Experiences was then performed over the 5 largest communities (Table 2) using GraphRAG integrated with GPT-3 5-turbo (temperature = 0) to digest content.

**Table 4.**
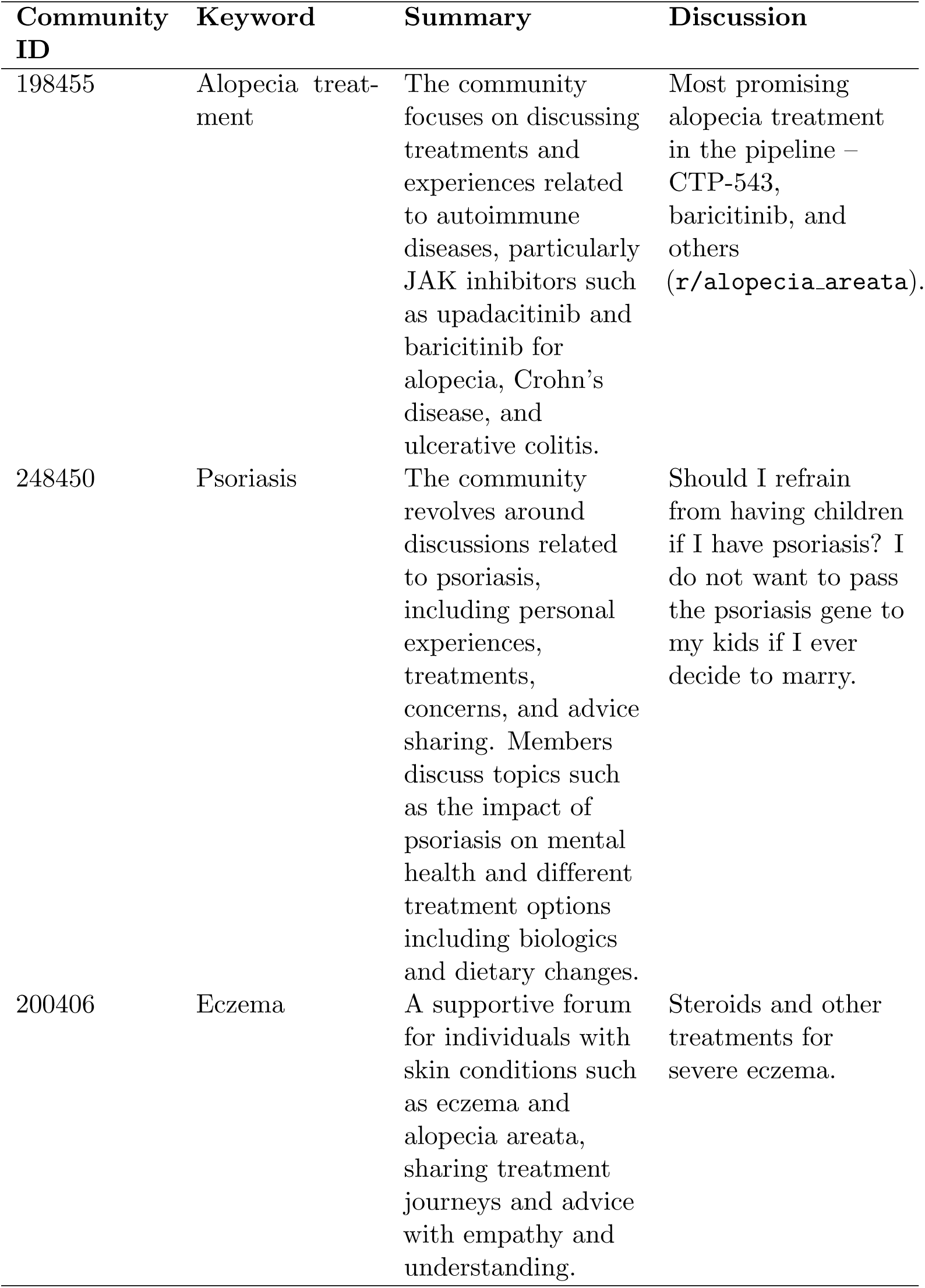
Labeled Communities and Their Characteristics in Reddit Discussions on JAK Inhibitors.

The degree distribution of the network followed a power law with an exponent of 2.11, confirmed by a Kolmogorov-Smirnov test (D = 0.47, p ¡ 0.0001) implying that a small number of high-connectivity users determined the flow of the discussion. Furthermore, PageRank analysis was applied to identify influential users with scores ranging from 0.752 to 0.961. For instance, Table (6) shows that the user with ID “1elda” from the “Eczema” community (PageRank = 0.961) and user with ID “e861b” from the “Alopecia areata” community (PageRank = 0.893) emerged as key-figures driving the conversations as they would initiate the discussion (i.e., posts) or amplify (i.e., replies) the threads.

**Table 6.**
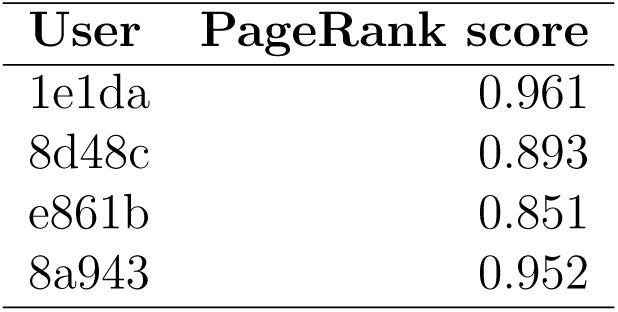
Patient-Reported Experiences Divergent from Current Medical Consensus in Reddit Communities.

The second query, “Return the 10 potential experiences divergent from current medical consensus in the top 5 communities detected regarding JAK inhibitors”, returned 50 sentences (10 per community) rated for exaggeration, evidence and contradictions to medical facts. These were randomly reviewed by a dermatologist after the large language model assigned them confidence scores between 0.6 and 0.9. Context was added to posts including eczema-related claims, where it said patients are “particularly vulnerable to unverified claims.” The model flagged 18.3% of posts and comments as potentially divergent from current evidence, indicating textual inconsistencies with established medical sources. These do not represent confirmed divergence from consensus.

**Table 7.** Patient-Reported Experiences Divergent from Current Medical Consensus in Reddit Communities.

| Post / Comment | Community | Divergent Patient Experience Explanation | Description |
| --- | --- | --- | --- |
| Quick rant here! Because it's getting annoying and frustrating that they always hype up the new drugs for eczema sufferers, for example the JAK inhibitors, but never want to discuss the adverse side effects. They always want to say it's mild and unrelated to therapy. Why do I feel like it's a lie? | Eczema | Alleges side effects of JAK inhibitors such as abrocitinib are downplayed or dismissed as unrelated, suggesting a cover-up. | The model flagged this sentence as potentially inconsistent with existing evidence (e.g., ORAL Surveillance), as it may mislead interpretation of risk-benefit balance. |
| I want to start with an overview of JAK inhibitors: no JAK inhibitor is yet approved for alopecia areata or its subtypes. | Alopecia areata | Claims no JAK inhibitors are approved for alopecia areata, implying lack of therapeutic potential despite later approvals. | The model flagged this statement as potentially inconsistent with current regulatory and clinical evidence. |
| I had patchy scalp alopecia which I regrew years ago using a home remedy. | Alopecia areata | Claims hair regrowth using a home remedy without scientific or clinical evidence. | Unverified recommendation that may encourage ineffective or misleading treatments over approved medical options. |
| I have signed up for a topical trial. My doctor mentioned this treatment could be better than Opzelura in terms of repigmentation and requires only once-daily application. I didn't want to participate in JAK inhibitor trials due to their serious side effects. | Vitiligo | States that JAK inhibitor trials involve serious side effects, presented as a generalised claim. | Side effects can occur, but severity varies; the phrasing may overgeneralise risk. |
| Baricitinib has not been shown to involve a shedding phase. | JAK inhibitor | Denies shedding phase for baricitinib, implying absolute safety relative to other therapies. | An overly certain safety claim; shedding can occur in some cases, potentially misleading patients about relative risks. |

Notable patient experiences divergent from current consensus were identified, demonstrating a range in the way experiences diverged from medical consensus. In one such case, a post by the user “1e1da” in alopecia areata stated that ”Quick rant here! Because It’s getting annoying and frustrating that they always hype up the new drugs for eczema sufferers”. This had a confidence score of 0.8, indicating its strong potential to mislead readers about subsequent approvals.

Divergent experiences were most frequently associated with exaggerated claims about side effects (e.g., framing all JAK inhibitor risks as severe or concealed) and promotion of unverified alternative treatments (e.g., home remedies), was identified by comparing user-generated content against established medical sources, including FDA labels, peer-reviewed trial data (e.g., ORAL Surveillance [67]), and expert clinical consensus (see Table 4). Claims were flagged as potentially divergent when they significantly diverged from this evidence, though we emphasize that user expressions often stem from personal experiences and uncertainty rather than intent to mislead. In contrast, efficacy-oriented discussions more closely reflected clinical trial outcomes and regulatory guidance.

Importantly, the use of TIMElinks, each of which carries a timestamp, allowed us to show temporal dynamics, including a notable increase in volume of discussion around the time 2022 FDA approval of baricitinib for alopecia areata as shown in figure(4), which illustrated the influence of regulatory events on community discourse. The GraphRAG pipeline was fine-tuned to mitigate false positives by 12% by raising the cosine similarity threshold to *>* 0.9. These patterns, primarily in alopecia and eczema communities, align with Vosoughi’s [59] findings on the rapid spread of false information, though the LLM classification results may reflect training data biases and the Reddit user base does not completely represent all patient demographics, so results should be interpreted with caution. This perspective exemplifies the value of using GraphRAG for real-time pharmacovigilance, while also emphasizing the need to enhance patient education to further connect health care professionals and the public.

**Fig 3.**
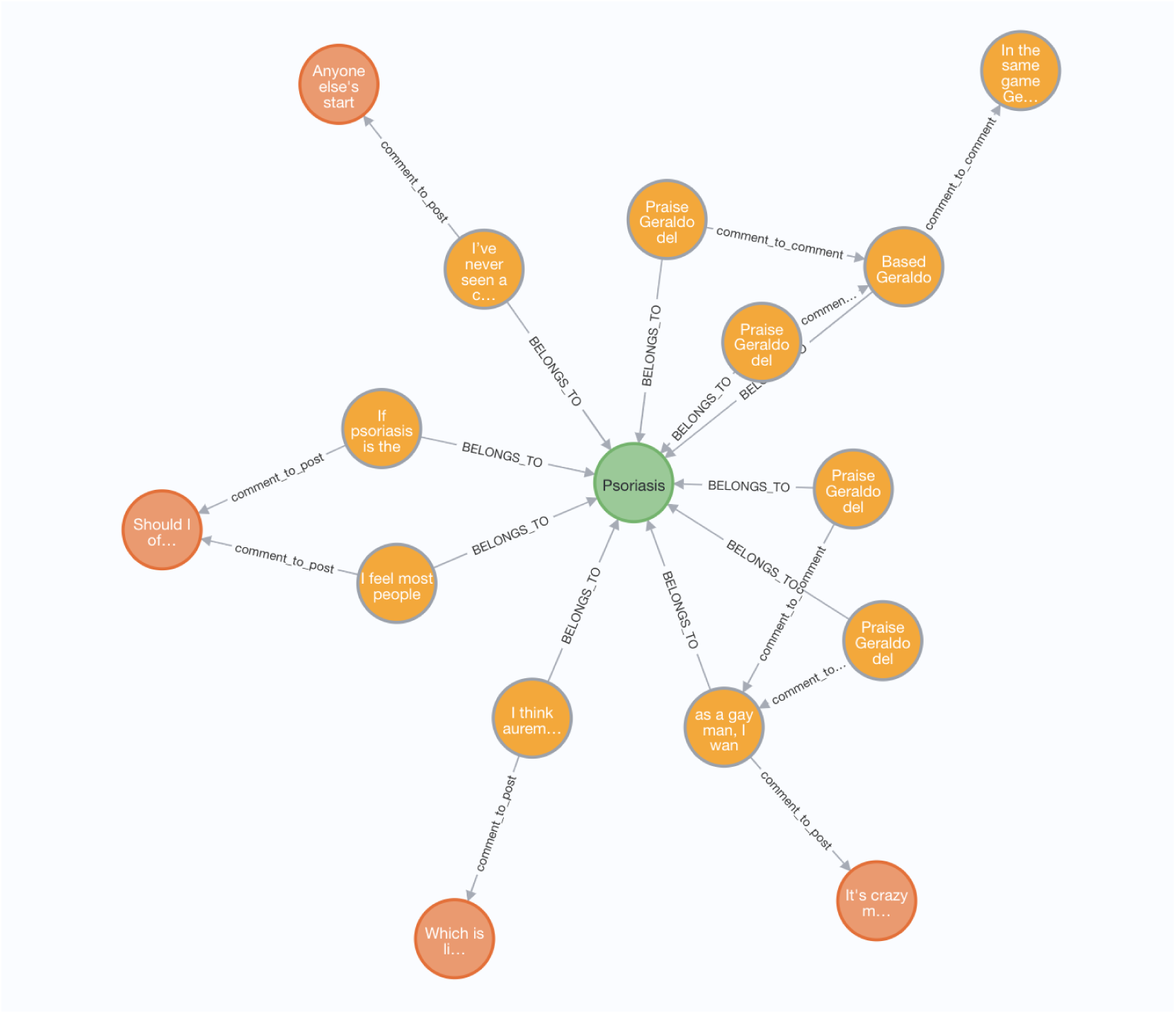
Sample Visualization of Psoriasis Community

**Fig 4.**
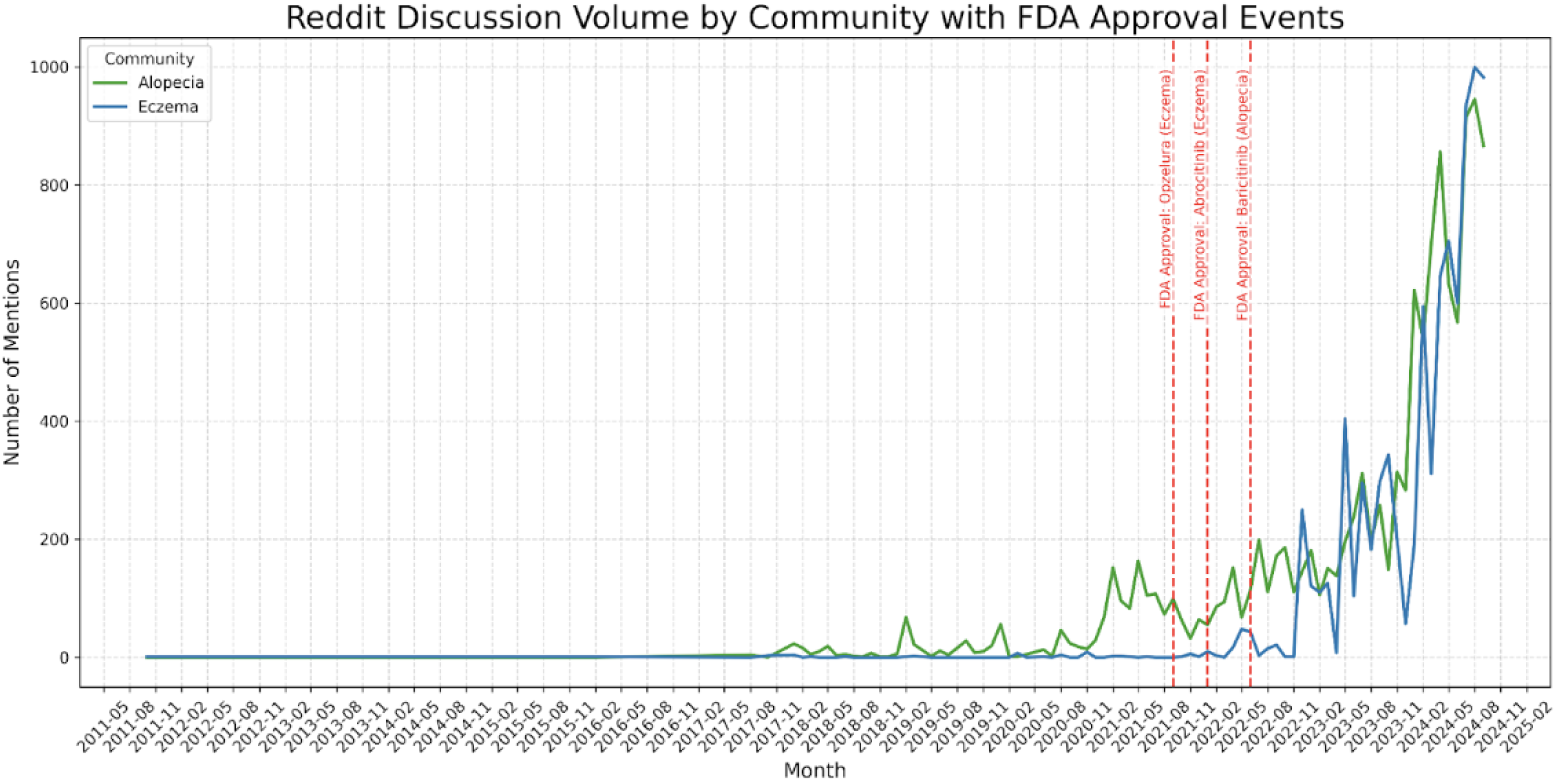
Reddit discussion volume for the Eczema and Alopecia communities from 2011 to 2024.

### 3.3 Critical Analysis of LLM Decision-Making

To address the need for transparency in how the LLM (GPT-3.5-turbo) prioritizes potentially divergent experiences within the dataset, a critical appraisal of its nature, bias, and limitations was undertaken. This model was fed structured prompts which included summarising and citing accepted medical advice (e.g., the FDA approvals and ORAL Surveillance study data) and asked to assess sentences’ divergence from the advice rather than make clinical decisions. For example, a post message like ”JAK inhibitors are a miracle cure with no side effects” from an eczema community would likely get a high score (e.g., 0.9) since the prompt asks the model to treat absolute claims of cure and absence of risk as potentially divergent from current evidence when compared to evidence showing mixed efficacy and known risks.

At implementation level, the model would score each post message by encoding the message into a text-to-vector form and returning per-sentence scores of how closely the sentences align with patterns described in the model prompt as potentially divergent from current evidence (e.g., exaggerated claims of efficacy, negation of known risks etc.). The scores and logical explanations were then provided sentence by sentence in the context of exploring which elements of a post had a greater influence on the model interpretation. Nevertheless, this approach has its problems and can be prone to errors.

An analysis of errors in a random sample of 10% of the dataset (22,145 items) indicated that about 12% of false positives (for instance, cases where personal stories or emotional reactions were flagged due to subjective language) and 8% of false negatives (for instance, instances of off-label promotion that did not clearly contradict guidance) occurred. Possible causes of bias might include the predominance of English-language training data, which would likely make the model focus on Western medical perspectives, and the risk of creating content that does not correspond to realistic facts.

Bias is also introduced by the Reddit dataset, as its user base is younger and more digitally engaged than the general patient population. For some conditions, this may lead to overrepresentation in the identified communities (for example, the proportion of alopecia-related posts may be higher compared with that for psoriasis), which in turn may amplify detection. We proposed mitigating these issues by iteratively refining the prompts, which included varying example statements, and validating a sample of 50 model-assigned scores with input from dermatologists, achieving 85% agreement between us and them on whether a sentence was potentially divergent from current evidence.

Significantly, the model does not make medical decisions; rather, it uses probabilistic heuristics that are indicative of pharmacovigilance research, and all the judgments indicating divergence from current consensus in this study are reliant on the evaluations of human experts. The next research phase may provide the interpretability tools designed to focus on which linguistic features specifically have an influence on the decisions of the model by using visual tools such as attention or saliency-based visualizations.

## 4 Discussions

This study provides an in-depth analysis of Reddit discussions surrounding JAK inhibitors, combining community detection, influencer analysis, and Analysis of Patient-Reported Experiences to advance real-time pharmacovigilance. A total of 337 communities with a modularity score of 0.9156 indicate that Reddit represents a clustered network of discussion on specific health conditions, such as alopecia and eczema. This aligns with prior research on online health discourse, which notes the formation of distinct conversational groups. The obtained scale-free network structure (power-law degree distribution, exponent = 2.11) and influential users (high PageRank scores) corroborate prior results indicating that a relatively small number of individuals drive a disproportionate amount of information flow through social networks, including for divergent patient experiences. However, this study builds upon these previous observations by leveraging GraphRAG and Neo4j to capture contextual nuances in JAK inhibitor discussions, allowing for the identification of particular divergent patient experiences examples that traditional keyword-matching approaches may miss. Addressing divergent patient experiences at the source, this study provides a novel framework for monitoring online health narratives in real-time with actionable insights for clinicians and regulators alike.

The identification of experiences divergent from current evidence highlights social media’s value as both a source of established patient insights and a discovery platform for emerging safety signals that may precede formal recognition, particularly in the context of JAK inhibitors. The proportion of model-flagged content was greater in alopecia and eczema communities (22.1% and 20.4%, respectively) than in psoriasis (15.7%). The 2021 ORAL Surveillance trial, for example, cast doubt on the safety profile of JAK inhibitors (eg, associations with major adverse cardiovascular events [67]), whereas the 2022 U.S. Food and Drug Administration approval of baricitinib for alopecia areata introduced new treatment pathways, resulting in a nuanced and sometimes contradictory information environment. These regulatory milestones were followed by spikes in divergent patient experiences, especially statements the model identified as potentially exaggerated (for example, presenting all risks associated with JAK inhibitors as severe) and unverified alternative treatments (for example, home remedies) were reported (Table 5). This pattern mirrors broader studies suggesting that health increases in content flagged as potentially inconsistent with evidence in response to clinical or regulatory announcements, but the spotlight on JAK inhibitors highlights distinctive challenges: patients in alopecia and eczema communities, with chronic conditions and few treatment options, may be especially susceptible to misleading narratives given the unmet needs and changing safety data. Comments from a dermatologist and the National Eczema Society were critical in making the outputs of the large language model clinically relevant and sensitive to these patients’ perspectives, so that divergent patient experiences that was flagged was contextualised by the lived experience of these communities.

The study’s primary contribution lies in its integration of Neo4j’s graph database with GraphRAG and large language models, surpassing traditional methods by capturing the interplay of community structures, influencer dynamics, and temporal trends in JAK inhibitor discussions. By focusing on influencers (Table 4) and temporal spikes (Fig 4), the study offers concrete insights into how and when experience sharings, so that it’s possible to gather targeted interventions, e.g., clinician-led education campaigns or regulatory monitoring, designed within the specific context of JAK inhibitors. For example, the increased rates of misleading narratives for JAK inhibitors after the ORAL Surveillance trial indicate that concerns about safety may be particularly potent in terms of perpetuating misleading information in these communities, a finding that can inform future risk communication of JAK inhibitors. The framework builds on the literature on health divergent patient experiences by providing a scalable, real-time pharmacovigilance mechanism, adaptable to other emerging therapies with complex safety profiles.

Future research could extend this approach to platforms like TikTok or include multilingual data or a broader perspective. Adjustments in model parameters might enhance accuracy further, and longitudinal studies could evaluate the lasting impact of divergent patient experiences following interventions. Connecting social media insight with clinical patient outcomes could anchor online narratives in real-world evidence. Working with regulatory agencies could leverage this framework for active safety monitoring.

## 5 Limitations

This study has several limitations. The LLM-generated divergence scores are probabilistic, prompt-guided estimates rather than validated clinical judgments, and therefore require expert interpretation that necessitates expert review for interpretation. Reddit users’ participation in this research is not an accurate reflection of the wider patient population.

Moreover, the discussions in English-language posts are quite limiting in terms of generalisability. Also, there have been more recent LLMs which have been released and become publicly available which may perform better. However, this is beyond the scope of this study and we leave it for future works.

Some conditions, such as alopecia and eczema, are overrepresented in the dataset, which may disproportionately influence detection rates within those communities. Although GraphRAG improved contextual retrieval, the system still fails to fully understand complicated or anecdotal statements. The possible drawbacks or limitations of the findings, as well as the applicability of the research to other pharmacovigilance settings, must be considered when drawing conclusions or making recommendations.

## 6 Conclusion

This study demonstrates the integration of graph-based, community detection, and large language models (LLMs) to characterize discourse in Reddit about Janus kinase (JAK) inhibitors, providing a novel perspective for real-time pharmacovigilance and analysis of patient-reported experiences in online health discussions. This perspective lies in the use of GraphRAG and Neo4j to analyze community structures, influencer dynamics, and temporal trends, providing a scalable, automated framework that surpasses traditional pharmacovigilance methods, such as manual reporting or post-marketing surveillance, enabling proactive identification of content that may be misleading or inconsistent with clinical guidance in online health discussions. We found that 18.3% of posts and comments about JAK inhibitors on Reddit, with rates of divergent patient experiences being higher in communities discussing alopecia treatments and eczema. Temporal analysis further showed peaks in divergent patient experiences following important regulatory events, including the 2022 FDA approval of baricitinib, emphasizing the responsiveness of online discourse to clinical developments.

## 7 Ethics Statement

This study analyzed on publicly available data from Reddit in accordance with Reddits Terms of Service and included no information that was deleted, private, or having any other restrictions. The username data was anonymized with irreversible hashing, and the researchers did not attempt to figure out the identity of the users. No user was contacted, thus no privacy issues or interventions of any online discussions made during the research took place, and it is considered that the research complies with the General Data Protection Regulation (GDPR). Results are shared in aggregate form in order to reduce the chances of harm or misinterpretation.

## Data Availability

In line with the terms and conditions of the Reddit online service, the authors do not have permission to share the data extracted from the social media platform.

## 8 Acknowledgments

We express our sincere gratitude to those who contributed to our research, and the institutions behind the work. We thank the Department of Computing and Mathematics at Manchester Metropolitan University for the computational resources and academic culture that facilitated this work. With a special mention to the dermatologists from Northern Care Alliance NHS Foundation Trust, UK for the expert review that contributed substantially to refining the clinical relevancy of our findings. We also thank the National Eczema Society for providing important context on patient vulnerabilities to unverified claims, enhancing our understanding of real-world impact. Thanks to the open-source communities behind tools like Neo4j, NetworkX, and asyncpraw, whose work made the data collection and analysis possible.

## Notes

### Competing Interest Statement

AA has received honoraria from UCB and Almirall, educational support from Almirall, Janssen and UCB to attend conferences, and has previously received funding from the UK North West MRC Scheme which is part-funded by Eli Lilly, UCB, Roche and Novartis.

### Funding Statement

Yes

### Author Declarations

Manchester Metropolitan University

